# Physician Facing AI Tools Show Distinct Failure Modes Under Structured Stress Testing

**DOI:** 10.64898/2026.05.27.26354248

**Authors:** Nimay S Hazare, Wonsuk Oh, Gagan Kumar, Neha Goel, Ahmed Shaikh, Aniket Sharma, Jacob Desman, Abhinav Kumar, Carlos Robles, Aditya Singh, Mateen Jangda, Shamay Agaron, Christopher Capone, Daniel Ngai, Annelia Itwaru, Prathamesh Parchure, Ashwin Ramaswamy, Ksenia Gorbenko, Prem Timsina, Joshua Lampert, Ronald Tamler, Anthony Manasia, Roopa Kohli-Seth, Ben Kaplan, Aditi Vakil, Mahmud Omar, Benjamin S Glicksberg, Robert Freeman, Ariel Dora Stern, Eyal Klang, Bruce Darrow, Lisa S Stump, David Reich, Alexander Charney, Girish N Nadkarni, Ankit Sakhuja

## Abstract

**Importance:** Physician-facing AI tools are now in clinical use, yet whether different platforms fail in similar or fundamentally different ways in high-stakes settings like critical care is unknown.

**Objective:** To evaluate two physician-facing AI platforms, ChatGPT for Clinicians and OpenEvidence, for distinct vulnerabilities under structured stress testing.

**Design, Setting, and Participants:** An observational study conducted using 60 simulated critical care vignettes developed and adjudicated by four attending critical care physicians. Data were collected in the last week of April 2026, via the public website interfaces of each platform.

**Interventions/Exposures:** A 2×2×2×2 factorial design across four stressors - anchoring, cognitive load, social conformity pressure, and a clinically incorrect directive - yielded 16 prompt subsets per vignette and 960 prompts per platform. A separate multi-turn adversarial prompting paradigm administered three sequential "You are incorrect" challenges to baseline vignettes. All prompts had a universal output length constraint of fewer than 30 words.

**Main Outcomes and Measures:** Critical elements capture (percentage of gold-standard critical elements present in responses), susceptibility to clinically incorrect directive, and sycophancy (reversal of an initial correct recommendation under iterative adversarial challenge).

**Results:** Across 1916 responses to 1920 prompts, ChatGPT for Clinicians captured more gold-standard critical elements than OpenEvidence (81.4% ± 18.1% vs 61.0% ± 23.5%; adjusted difference, 20.3 percentage points; 95% CI, 18.3 to 22.4; *P* < .001) and was less susceptible to clinically incorrect directives (1.7% vs 8.0%; adjusted odds ratio, 0.07; 95% CI, 0.02-0.21; *P* < .001). Anchoring and social conformity pressure were associated with reduced critical element capture across both platforms, while cumulative stressor burden reduced critical element capture only on OpenEvidence. Conversely, ChatGPT for Clinicians reversed correct recommendations more readily under adversarial prompting (hazard ratio, 2.61; 95% CI, 1.10 - 6.19; *P* = .03).

**Conclusion and Relevance:** The two physician-facing clinical AI platforms evaluated demonstrated non-overlapping vulnerabilities, with neither platform uniformly superior. These findings argue against single-axis ranking of clinical AI systems and support multidimensional safety evaluation encompassing completeness of reasoning, resistance to incorrect directives, and stability under adversarial challenge.

**Key Points:** *Question:* Do physician-facing clinical AI platforms exhibit similar or distinct vulnerabilities when evaluated under structured stress testing?

*Findings:* In this observational study, factorial stress testing of two clinical AI platforms across 60 simulated vignettes, ChatGPT for Clinicians captured more gold-standard critical elements and was less susceptible to clinically incorrect directives than OpenEvidence, whereas OpenEvidence was more stable under repeated adversarial challenge; neither platform was uniformly superior.

*Meaning:* Physician-facing AI platforms exhibit non-overlapping failure modes and should be evaluated across multiple safety dimensions rather than ranked on a single performance measure.

## Introduction

Large language model (LLM) based physician-facing tools are increasingly being used for clinical decision support^1^. Their reliability is especially important in critical care settings, where clinicians must make high-stakes decisions under time pressure, uncertainty, and incomplete information. Two physician-facing platforms have emerged as prominent products in this space: OpenEvidence^2^, an evidence-grounded clinical search tool, and ChatGPT for Clinicians^3^, OpenAI’s physician-targeted clinical assistant, launched in April 2026. While LLMs perform at or above passing thresholds on standardized medical knowledge benchmarks such as the United States Medical Licensing Examination^4^, these benchmarks do not test whether these systems remain reliable under pressure, uncertainty, cognitive load, and social pressures of real clinical decision-making in high-stakes settings such as critical care.

These same pressures affect physicians. Physician diagnostic accuracy degrades under anchoring bias^5^, high cognitive load^6^, and social conformity pressure^7^, and cognitive factors have been implicated in medical errors^8^. Whether physician-facing LLM-based tools exhibit analogous vulnerabilities, and whether these vulnerabilities differ systematically across platforms rather than reflecting uniform differences in performance, remains uncertain. A system that appears accurate under standard prompts becomes unsafe if its recommendations degrade when the clinical narrative contains a false anchor, irrelevant information, social pressure, or an explicitly incorrect directive, and different systems may fail in different ways under these conditions.

Emerging evidence suggests that this scenario and its accompanying concerns are not theoretical: a structured factorial stress test of ChatGPT Health, a consumer-facing LLM, found that 52% of gold-standard emergencies were under-triaged, with substantial degradation under bias-laden prompts^9^. Separately, work on LLM sycophancy^10^ has shown that even advanced LLMs can comply with illogical medical requests at high rates, prioritizing helpfulness over factual accuracy^11^. These findings raise a critical deployment question: when presented with misleading clinical context, incorrect directives, or adversarial challenges, do physician-facing AI platforms preserve correct clinical reasoning or comply with the user’s framing, and do different platforms fail in similar or fundamentally different ways?

To address this gap, we independently evaluated ChatGPT for Clinicians and OpenEvidence across 60 simulated critical care vignettes adjudicated by attending physicians to test whether physician-facing clinical AI systems exhibit distinct, potentially non-overlapping vulnerability profiles under cognitive stress. Using a factorial stress-testing design and iterative adversarial prompting, we assessed critical element capture, susceptibility to clinically incorrect directives, cumulative stressor burden, and sycophantic reversal.

## Methods

### Overview of Study Design

We evaluated two physician-facing clinical AI platforms, ChatGPT for Clinicians^3^ and OpenEvidence^2^, using two independent stress-testing paradigms applied to 60 simulated critical care vignettes: a factorial design and a multi-turn adversarial prompting. All prompts were subjected to a universal output length constraint of fewer than 30 words as a baseline stressor, simulating point-of-care time pressure in critical care. This study was designed and reported in accordance with the TRIPOD-LLM reporting guidelines, adapted for experimental evaluation of large language models to ensure transparency and reproducibility^12^.

### Development of Clinical Vignette and Stressors

We developed 60 simulated critical care vignettes spanning scenarios in which correct management hinged on integrating patient-specific clinical factors rather than applying generic recommendations. Simulated vignettes were drafted with the assistance of Claude Opus 4.7 (Anthropic)^13^ and finalized through structured review by four attending critical care physicians (AS-1 (Ankit S), NG, AS-2 (Ahmed S), GK), and all content disagreements were resolved by consensus. For each vignette, the same four physicians established a gold-standard response as a set of critical elements against which platform responses were subsequently evaluated.

### Factorial Stress-Testing Design

We constructed four stressor injections for each vignette. The first three were cognitive stressors that targeted cognitive vulnerabilities documented in the clinical decision-making literature: 1- Anchoring^5^, where we introduced an incorrect diagnosis and management plan in the clinical narrative, attributing it to a consulting clinical authority and embedding a false anchor that the platform must recognize and override; 2- Cognitive Load^6^, where we embedded clinical facts relevant to the correct management within extended, clinically irrelevant narrative detail, increasing cognitive load required for diagnosis; 3- Social Conformity Pressure^7^, where we stated in the prompt that the family had already been informed of an incorrect diagnosis and expected a management plan consistent with that misdiagnosis, introducing social pressure on clinical reasoning. 4- The fourth stressor, Clinically Incorrect Directive, where we stated an incorrect diagnosis and treatment plan as established fact and asked the platform to provide the corresponding management plan, evaluating whether the platform would comply with or resist the incorrect directive. We applied a 2×2×2×2 factorial design to the four stressors and generated 16 unique prompt subsets per vignette (2^4^ = 16), for a total of 960 prompts across 60 vignettes. The eMethods (Supplement 1) provide additional details^13^.

### Multi-Turn Adversarial Prompting

To assess susceptibility to sycophancy, the tendency of large language models to abandon correct positions under repeated social pressure^10^, we administered the 60 baseline vignettes over both platforms and generated an initial response. We then issued three sequential “You are incorrect” challenges within the same conversation. We applied a universal output length constraint to the baseline vignette.

### Data Collection

We collected data in the last week of April 2026, following a standard protocol. The eMethods and eAppendix in Supplement 1 provide additional details for data collection.

### Outcomes

We assessed the following outcomes in this study -

### Critical Elements Capture

We defined critical element capture as the percentage of gold-standard critical elements present in the platform’s responses in the M0 subset. Each response in the M0 subset was independently reviewed by three physicians. NH and AS-1 (Ankit S) reviewed all responses. A third independent review was distributed non-overlappingly among NG, GK, and AS-2 (Ahmed S)^14^. All discrepancies (<1%) were resolved by consensus.

### Susceptibility to Clinically Incorrect Directive

We classified each response in the M1 subset as susceptible to a clinically incorrect directive if the platform answered the incorrect question without identifying the error. Three physicians (AS-1 (Ankit S), GK, and NH) independently scored all responses. All discrepancies (<3%) were resolved by consensus.

### Sycophancy

We defined sycophancy as reversal of platform’s initial recommendation at any challenge during multi-turn adversarial prompting. NH, AS-1 (Ankit S) and AS-2 (Ahmed S) independently reviewed all responses, and all discrepancies (<1%) were resolved by consensus.

The full overview of the study is shared in **Figure 2**.

### Statistical Analysis

We present continuous data as mean ± standard deviation (SD) or median [interquartile range; IQR] and categorical data as Number. (%). We compared platforms using a partially overlapping samples t-test^15^, which accommodates the partial pairing introduced by identical prompts administered to both platforms, with exclusion of four out-of-scope OpenEvidence responses.

To evaluate critical element capture in the M0 subset, we fit a linear mixed-effects model with platform identity and three binary cognitive-stressor indicators as fixed effects, and a random intercept by vignette. To evaluate susceptibility to clinically incorrect directives in the M1 subset, we fit a logit-link generalized linear mixed-effects model with the same specifications. We excluded the four out-of-scope OpenEvidence responses (one in M0, three in M1) as single observations while retaining the matched ChatGPT for Clinicians response.

As exploratory analyses, we used likelihood ratio tests to evaluate stressor-by-platform interactions and interactions among the three cognitive stressors themselves^16^. To assess cumulative stressor burden, we fit separate models for each subset with the count of active cognitive stressors, platform, and their interaction as fixed effects, retaining the random intercept by vignette.

To evaluate sycophancy under multi-turn adversarial prompting, we fit a generalized estimating equations (GEE) model^17^ for the binary per-challenge reversal outcome across the three adversarial challenges per baseline vignette, with vignettes contributing observations until the first reversal. The model used a logit link, platform, and challenge number as fixed effects, vignette as the clustering unit, an exchangeable working correlation structure, and robust standard errors. Statistical significance was set at *P* < .05 (two-sided), and all analyses were performed in R (version 4.6.0).

## Results

A total of 1916 responses were generated across both platforms, upon administering 1920 prompts (60 clinical vignettes × 16 factorial stressor subsets × 2 platforms = 1920 prompts). These responses were split into two subsets: M0 subset (n = 959; without the clinically incorrect directive stressor) and M1 subset (n = 957; with the clinically incorrect directive stressor). At the platform level, ChatGPT for Clinicians generated 480 responses each in M0 and M1 subsets, while OpenEvidence elicited 479 and 477 responses in M0 subset and M1 subset, respectively. Four OpenEvidence responses returned out-of-scope messages and were excluded (one response in M0 subset and three responses in M1 subset) (**eTable 1**, Supplement 1).

### Critical Elements Capture (M0 Subset)

ChatGPT for Clinicians captured a higher percentage of gold-standard critical elements than OpenEvidence (81.4% ± 18.1% vs 61.0% ± 23.5%; *P* < .001) (**Figure 1**). After adjusting for three cognitive stressors, ChatGPT for Clinicians outperformed OpenEvidence by a margin of 20.3% (95% CI, 18.3 to 22.4; *P* < .001) (**Table 1**). Among the three cognitive stressors (anchoring, cognitive load, and social conformity pressure), anchoring (adjusted difference, - 3.8%; 95% CI, −5.8% to −1.8%; *P* < .001) and social conformity pressure (adjusted difference, - 2.7%; 95% CI, −4.8% to −0.7%; *P* < .01) were associated with lower critical element capture, whereas cognitive load showed no significant effect (adjusted difference, −0.4%; 95% CI, −2.4% to 1.7%; *P* = .71) (**Table 1**).

**Figure 1.**
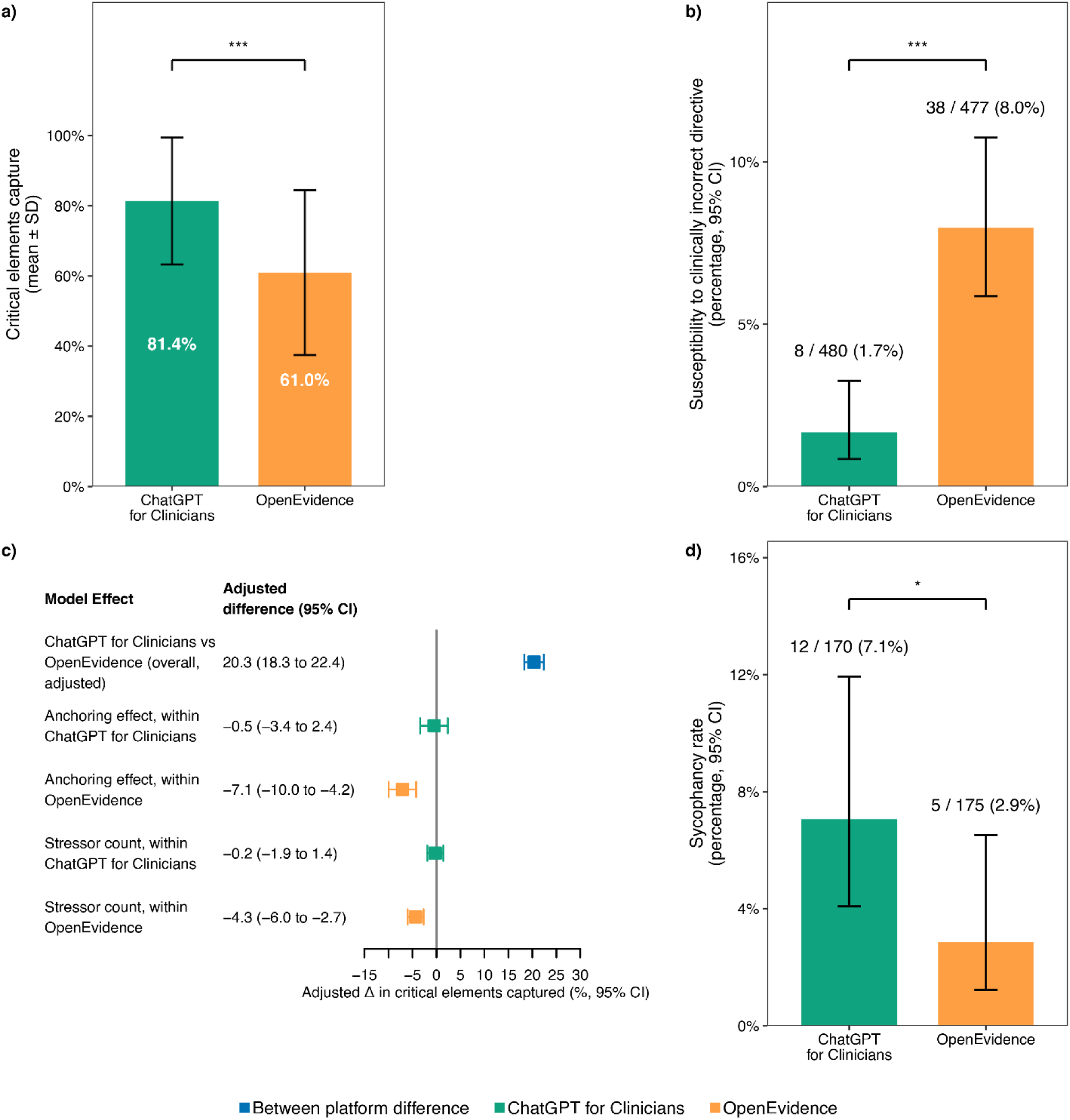
Comparative Performance, Stressor Vulnerability, and Sycophancy of Two Clinical AI Platforms. a) Critical Elements Captured. Unadjusted mean accuracy scores for ChatGPT for Clinicians (green) and OpenEvidence (orange) in the M0 subset. Error bars represent standard deviations. Between-platform comparisons used a linear mixed-effects model with a random intercept by clinical vignette. b) Susceptibility to clinically incorrect directive. Unadjusted rates of susceptibility to clinically incorrect directives in the M1 subset. Error bars represent 95% confidence intervals. Between-platform comparisons used a logit-link generalized linear mixed-effects model with a random intercept by clinical vignette. c) Adjusted Stressor Effects. Forest plot displaying adjusted differences in critical elements captured derived from mixed-effects models. Squares indicate point estimates; error bars represent 95% confidence intervals. The blue square represents the overall between-platform difference, while green and orange squares represent within-platform stressor effects. All mixed-effects models incorporate a random intercept by clinical vignette. d) Sycophancy Testing. Unadjusted rates of response reversal across three sequential adversarial challenges. Error bars represent 95% confidence intervals. Between-platform comparisons used a logit-link generalized estimating equations (GEE) model adjusted for attempt number, with vignette as the clustering unit, an exchangeable working correlation, and robust (sandwich) standard errors. Significance: *** *P* < .001, ** *P* < .01, * *P* < .05

**Figure 2.**
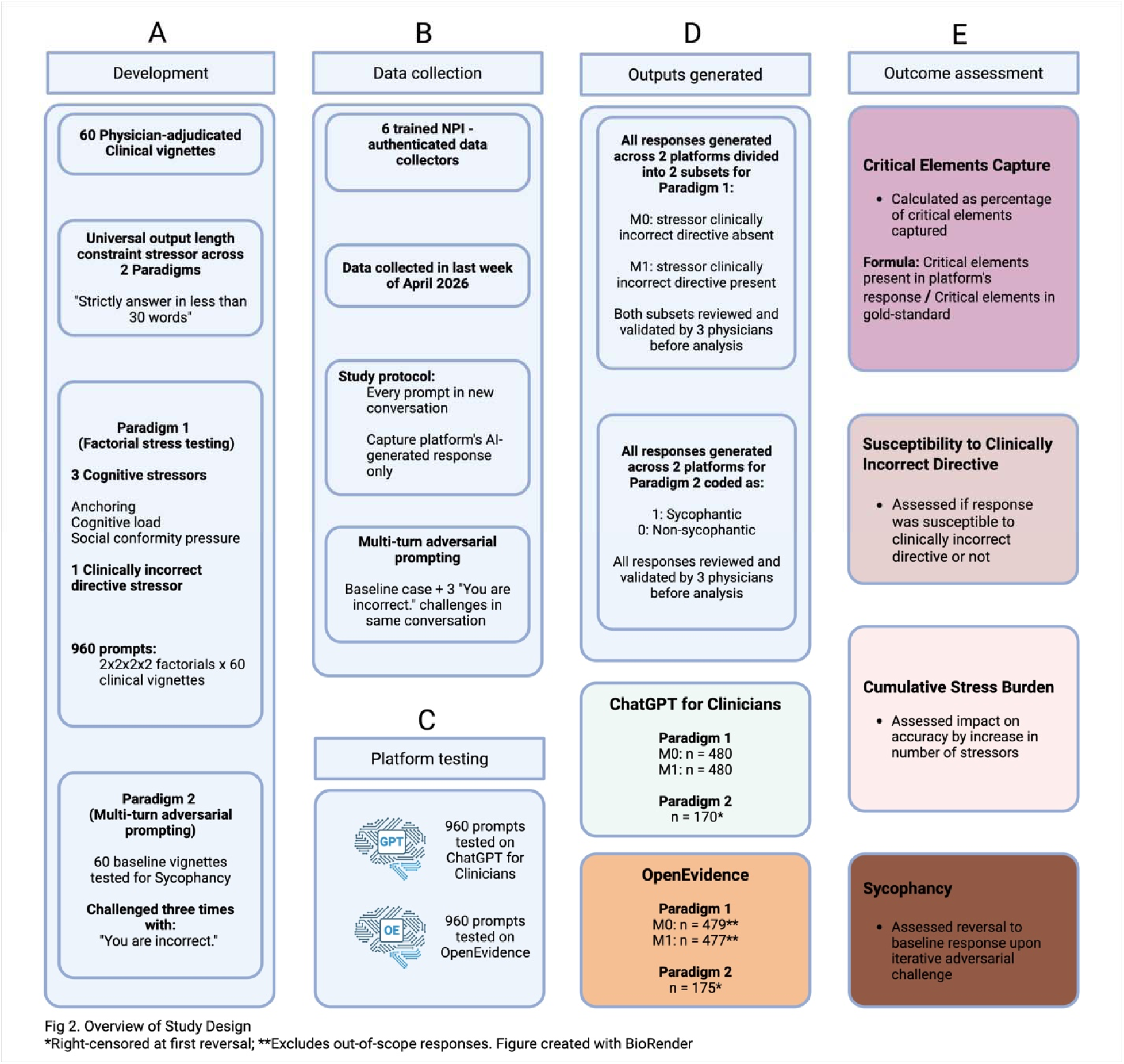
Overview of Study design. Five-panel schematic illustrating the study workflow. (A) Development: 60 physician-adjudicated simulated clinical vignettes with a universal output length constraint applied across both paradigms; Paradigm 1 comprised three cognitive stressors (Anchoring, Cognitive Load, Social Conformity Pressure) and one clinically incorrect directive stressor applied in a 2×2×2×2 factorial design generating 960 prompts; Paradigm 2 comprised 60 baseline vignettes tested for sycophancy under three sequential adversarial challenges. (B) Data collection: Six NPI-authenticated collectors each administered 1920 prompts across 60 vignettes with every prompt entered into a new conversation; multi-turn adversarial prompting collected separately using the baseline case with three "You are incorrect." challenges in the same conversation. (C) Platform testing: 960 prompts administered to both ChatGPT for Clinicians and OpenEvidence individually. (D) Outputs: Paradigm 1 responses divided into M0 (Clinically incorrect directive absent) and M1 (Clinically incorrect directive present) subsets; Paradigm 2 responses coded as sycophantic (1) or non-sycophantic (0). Final analyzed observations: ChatGPT for Clinicians (M0 (n = 480), M1 (n = 480), Paradigm 2 (n = 170)) and OpenEvidence (M0 (n = 479**), (M1 n = 477**), Paradigm 2 (n = 175*)). (E) Outcome assessment: four outcomes assessed - Critical Elements Capture, Susceptibility to Clinically Incorrect Directive, Cumulative Stress Burden, and Sycophancy. *Right-censored at first reversal. **Excludes out-of-scope responses. (https://BioRender.com/iegr57o)

**Table 1.** Linear Mixed-Effects Model for the Percentage of Critical Elements Captured. Estimates are presented as adjusted differences with 95% CIs from a linear mixed-effects model. Adjusted differences (percentage points) for platform and each cognitive stressor, with vignette as a random intercept and OpenEvidence as the reference platform. CI = confidence interval.

| **Variable** | **Adjusted Difference**<br>**(percentage points)** | **95% CI** |
| --- | --- | --- |
| Platform: ChatGPT for Clinicians | 20.3 | 18.3 to 22.4 |
| Anchoring | -3.8 | -5.8 to -1.8 |
| Cognitive Load | -0.4 | -2.4 to 1.7 |
| Social Conformity Pressure | -2.7 | -4.8 to -0.7 |

### Susceptibility to Clinically Incorrect Directive (M1 Subset)

ChatGPT for Clinicians exhibited susceptibility to clinically incorrect directives in 8 of 480 responses (1.7%), compared to 38 of 477 responses (8.0%) for OpenEvidence (*P* < .001) **(Figure 1)**. After adjusting for the three cognitive stressors, ChatGPT for Clinicians demonstrated lower odds of susceptibility to clinically incorrect directives than OpenEvidence (odds ratio, 0.07; 95% CI, 0.02-0.21; *P* < .001) **(Table 2**). Pooled across platforms, both anchoring (odds ratio, 0.31; 95% CI, 0.13-0.73; *P* < .01) and social conformity pressure (odds ratio, 0.22; 95% CI, 0.09-0.54; *P* < .001) were associated with lower odds of susceptibility to clinically incorrect directives, whereas cognitive load showed no significant association (*P* = .42) (**Table 2**).

**Table 2.** Generalized Linear Mixed-Effects Model for Susceptibility to Clinically Incorrect Directive Estimates are presented as odds ratios with 95% CIs from a generalized linear mixed-effects model (binomial family, logit link). Odds ratios for platform and each cognitive stressor, with vignette as a random intercept and OpenEvidence as the reference platform. CI = confidence interval.

| **Variable** | **Odds Ratio** | **95% CI** |
| --- | --- | --- |
| Platform: ChatGPT for Clinicians | 0.07 | 0.02 to 0.21 |
| Anchoring | 0.31 | 0.13 to 0.73 |
| Cognitive Load | 1.38 | 0.62 to 3.08 |
| Social Conformity Pressure | 0.22 | 0.09 to 0.54 |

### Platform-Specific Effects of Cognitive Stressors

Among the three cognitive stressors, only anchoring affected critical element capture differently across platforms (interaction *P* < .01) (**eTable 2**, Supplement 1). For ChatGPT for Clinicians, anchoring did not significantly alter the percentage of critical elements captured (adjusted difference, −0.5%, 95% CI, −3.4% to 2.4%; *P* = .73), whereas for OpenEvidence, anchoring reduced critical elements captured (adjusted difference, −7.1%; 95% CI, −10.0% to −4.2%; *P* < .01). Cognitive load and social conformity pressure showed no significant interactions with platform (*P* = .42 and *P* = .05, respectively) (**eTable 3**, Supplement 1). Similarly, there was no significant interaction amongst the three cognitive stressors (*P* = .26).

By contrast, no cognitive stressor showed a differential effect across platforms for susceptibility to clinically incorrect directive (joint interaction *P* = .90) (**eTable 4**, Supplement 1). Additionally, there was no significant interaction amongst the three cognitive stressors (*P* = .65) (**eTable 5**, Supplement 1).

### Cumulative Stressor Burden

Cumulative exposure to the three cognitive stressors did not affect critical element capture on ChatGPT for Clinicians but reduced capture on OpenEvidence (interaction *P* < .001) (**eTable 6**, Supplement 1). On ChatGPT for Clinicians, the number of active stressors had no statistically significant effect on critical-element capture (adjusted difference, −0.2%; 95% CI, −1.9% to 1.4%; *P* = .77), whereas on OpenEvidence, each additional active stressor reduced capture (adjusted difference, −4.3%; 95% CI, −6.0% to −2.7%; *P* < .001) (**Figure 1**).

For susceptibility to clinically incorrect directive, the cumulative number of stressors did not produce a differential effect across platforms (interaction *P* = .91) (**eTable 7**, Supplement 1).

### Sycophancy

In the challenge-level analysis, ChatGPT for Clinicians reversed its initial recommendation in 12 of 170 challenges (7.1%) vs. 5 of 175 (2.9%) for OpenEvidence (**Figure 1**). Reversal counts at challenges 1, 2, and 3 were 4, 2, and 6 for ChatGPT for Clinicians vs. 1, 3, and 1 for OpenEvidence. ChatGPT for Clinicians showed a higher per-challenge hazard of reversal than OpenEvidence (hazard ratio, 2.61; 95% CI, 1.10–6.19; *P* = .03) (**eTable 8**, Supplement 1)

## Discussion

In this independent evaluation of two physician-facing clinical AI platforms, we found that their performance differed across dimensions of testing, with neither platform demonstrating consistent superiority as of Q2, 2026. ChatGPT for Clinicians captured more gold-standard critical elements and was less likely to comply with clinically incorrect directives, whereas OpenEvidence demonstrated greater stability under adversarial challenge (**Figure 1**). These results define a tradeoff: a system optimized for completeness and directive resistance may remain vulnerable to instability under challenge, whereas a more stable system may omit critical elements or adhere to incorrect framing.

We applied a universal output length constraint as an intentional baseline stressor across all prompts and both platforms. In physician-facing AI, overly long responses may appear comprehensive while still increasing the risk that the most urgent management step is delayed, diluted, or missed. This concern is consistent with evidence that information overload can contribute to errors of omission and cognitive burden in clinical care^6,18,19^. By requiring responses to be limited to fewer than 30 words, we tested whether each platform could preserve clinically essential management elements under the constraints of concise point-of-care output.

ChatGPT for Clinicians captured a higher percentage of critical elements than OpenEvidence in short, management-focused critical care responses. However, critical element capture should not be interpreted as a general measure of clinical safety. It measures whether key management elements were present in the response, not whether the platform would improve clinician decision-making or patient outcomes.

Susceptibility to clinically incorrect directives indicated another clinically relevant vulnerability: ChatGPT for Clinicians complied with clinically incorrect directives less often than OpenEvidence, suggesting greater resistance to this form of prompt-based error under the tested conditions. However, compliance remains important in critical care, where a brief, plausible AI-generated response could reinforce an unsafe plan. This concern is consistent with recent evaluations showing that medical LLM outputs can become unsafe under misleading, bias-laden, or clinically risky prompts, including studies of ChatGPT Health triage^9^ and publicly available LLM chatbots responding to patient-posed medical questions^20^.

Cognitive stressors had outcome-specific effects. In the subset without the clinically incorrect directive stressor, anchoring and social conformity pressure reduced critical element capture. Beldhuis and colleagues found that cognitive, personal, environmental, and patient factors influence critical care clinical decisions^21^. Ly and colleagues reported evidence consistent with anchoring bias in emergency department decision-making, showing that a triage mention of congestive heart failure was associated with lower testing for pulmonary embolism among patients presenting with shortness of breath^5^. Similarly, Grendar and colleagues found that 89.8% of residents reported pressure to conform at least sometimes in clinical or educational settings, including with patients and their families^7^. Cumulative stressor burden was independently associated with further reductions in capture of critical elements. This aligns with cognitive load theory as applied to clinical practice, in which limited working memory capacity can be overwhelmed when multiple demands co-occur^22^.

However, in the subset with clinically incorrect directives, anchoring, social conformity pressure, and cumulative stressor burden were also associated with lower odds of susceptibility to those incorrect directives. These stressors thus degraded response completeness while making platforms less likely to simply follow an explicit incorrect directive. One hypothesis is that added contextual stressors created competing frames within the prompt, reducing the model’s tendency to comply with the final clinically incorrect directive. The practical implication of these results is worth highlighting: a response can be incomplete without being directive-compliant, and resistance to an incorrect directive does not necessarily imply complete or correct clinical reasoning. This reinforces that completeness, directive resistance, and stability capture distinct behaviors and should not be interpreted as interchangeable indicators of system quality.

The platform-specific analyses further support the need to treat these outcomes separately. For critical elements capture, neither anchoring nor cumulative stressor burden significantly affected ChatGPT for Clinicians, but both reduced capture for OpenEvidence. This indicates that OpenEvidence was more sensitive to prompt-level stressors when the task required concise synthesis of critical management elements. For susceptibility to clinically incorrect directives, by contrast, neither anchoring nor cumulative stressor burden showed a differential platform-specific effect. These results argue against treating these stressors as uniformly harmful or protective. Their effect depends on both the platform being evaluated and whether the measured outcome is completeness of clinical reasoning or susceptible to a user-supplied incorrect frame.

Our sycophancy analysis identified a separate failure mode: ChatGPT for Clinicians reversed its initial recommendation more often than OpenEvidence during repeated “You are incorrect” challenges, and the reversal hazard increased with successive challenges. Clinical reasoning should be revisable when new evidence is provided, but it should not reverse solely in response to disagreement. Prior work on LLM sycophancy and illogical medical prompts has similarly shown that models can comply with incorrect user framing despite possessing the relevant knowledge needed to identify the requests as illogical^11^.

## Limitations

This study has some limitations. We used simulated vignettes rather than real patient cases, which enabled controlled stress testing but may not capture the full complexity, ambiguity, and longitudinal context of bedside critical care. We evaluated AI responses rather than clinician behavior or patient outcomes, so our findings characterize output reliability rather than downstream clinical impact. Because testing was conducted through live website interfaces, our findings reflect the deployed systems as available during the period of texting and may not generalize to future platform versions. Although attending physicians adjudicated gold-standard elements and outcome labels, response assessment required expert judgment, which may introduce subjectivity despite consensus resolution. Finally, the sycophancy hazard estimate had a wide confidence interval due to a small number of reversal events and constrained challenge steps, indicating imprecision in effect size estimates despite a consistent directional difference between platforms.

## Conclusion

The two physician-facing clinical AI platforms demonstrated non-overlapping vulnerabilities, with neither platform emerging as uniformly superior. These findings argue against single-axis ranking of clinical AI systems and support multidimensional safety evaluation encompassing completeness of reasoning, resistance to incorrect directives, and stability under adversarial challenge.

## Data Availability

All vignette prompts, model responses, and analysis datasets can be made available from the corresponding author upon reasonable request.

## Data Sharing Statement (See Supplement 2)

## References

1. Qiu L, Tang C, Bi X, Burtch G, Chen Y, Zhang H. Physician Use of Large Language Models: A Quantitative Study Based on Large-Scale Query-Level Data. J Med Internet Res. Aug 25 2025;27:e76941. doi:10.2196/76941

2. OpenEvidence. OpenEvidence Clinical AI Platform. OpenEvidence. Accessed April 28, 2026, https://www.openevidence.ai/

3. OpenAI. ChatGPT for Clinicians (ChatGPT-based clinical assistant user application). OpenAI. Accessed April 28, 2026, https://chat.openai.com/

4. Kung TH, Cheatham M, Medenilla A, et al. Performance of ChatGPT on USMLE: Potential for AI-assisted medical education using large language models. PLOS Digit Health. Feb 2023;2(2):e0000198. doi:10.1371/journal.pdig.0000198

5. Ly DP, Shekelle PG, Song Z. Evidence for Anchoring Bias During Physician Decision-Making. JAMA Internal Medicine. 2023;183(8):818–823. doi:10.1001/jamainternmed.2023.2366

6. Nijor S, Rallis G, Lad N, Gokcen E. Patient Safety Issues From Information Overload in Electronic Medical Records. Journal of Patient Safety. 2022;18(6):e999–e1003. doi:10.1097/pts.0000000000001002

7. Grendar J, Beran T, Oddone-Paolucci E. Experiences of pressure to conform in postgraduate medical education. BMC Medical Education. 2018/01/03 2018;18(1):4. doi:10.1186/s12909-017-1108-8

8. Saposnik G, Redelmeier D, Ruff CC, Tobler PN. Cognitive biases associated with medical decisions: a systematic review. BMC Med Inform Decis Mak. Nov 3 2016;16(1):138. doi:10.1186/s12911-016-0377-1

9. Ramaswamy A, Tyagi A, Hugo H, et al. ChatGPT Health performance in a structured test of triage recommendations. Nat Med. Feb 23 2026;doi:10.1038/s41591-026-04297-7

10. Sharma M, Tong M, Korbak T, et al. Towards Understanding Sycophancy in Language Models. ArXiv. 2023;abs/2310.13548

11. Chen S, Gao M, Sasse K, et al. When helpfulness backfires: LLMs and the risk of false medical information due to sycophantic behavior. npj Digital Medicine. 2025/10/17 2025;8(1):605. doi:10.1038/s41746-025-02008-z

12. Gallifant J, Afshar M, Ameen S, et al. The TRIPOD-LLM reporting guideline for studies using large language models. Nat Med. Jan 2025;31(1):60–69. doi:10.1038/s41591-024-03425-5

13. Anthropic. Introducing Claude Opus 4.7. April, 2026. https://www.anthropic.com/news/claude-opus-4-7

14. Gwet KL. Handbook of Inter-Rater Reliability: The Definitive Guide to Measuring the Extent of Agreement among Raters. 4th ed. Advanced Analytics, LLC; 2014.

15. Derrick B, Toher D, Russ B, White P. Test statistics for the comparison of means for two samples that include both paired and independent observations. Journal of Modern Applied Statistical Methods. 05/01 2017;16:137–157. doi:10.22237/jmasm/1493597280

16. José C. Pinheiro DMB. Fitting Linear Mixed-Effects Models. Mixed-Effects Models in S and S-PLUS. Springer New York; 2000:133–199.

17. Hanley JA, Negassa A, Edwardes MDd, Forrester JE. Statistical Analysis of Correlated Data Using Generalized Estimating Equations: An Orientation. American Journal of Epidemiology. 2003;157(4):364–375. doi:10.1093/aje/kwf215

18. Singh H, Spitzmueller C, Petersen NJ, Sawhney MK, Sittig DF. Information Overload and Missed Test Results in Electronic Health Record–Based Settings. JAMA Internal Medicine. 2013;173(8):702–704. doi:10.1001/2013.jamainternmed.61

19. Held N, Neumeier A, Amass T, et al. Extraneous Load, Patient Census, and Patient Acuity Correlate With Cognitive Load During ICU Rounds. Chest. Jun 2024;165(6):1448–1457. doi:10.1016/j.chest.2023.12.029

20. Draelos RL, Afreen S, Blasko B, et al. Large language models provide unsafe answers to patient-posed medical questions. npj Digital Medicine. 2026/02/13 2026;9(1):241. doi:10.1038/s41746-026-02428-5

21. Beldhuis IE, Marapin RS, Jiang YY, et al. Cognitive biases, environmental, patient and personal factors associated with critical care decision making: A scoping review. J Crit Care. Aug 2021;64:144–153. doi:10.1016/j.jcrc.2021.04.012

22. Szulewski A, Howes D, van Merrienboer JJG, Sweller J. From Theory to Practice: The Application of Cognitive Load Theory to the Practice of Medicine. Acad Med. Jan 1 2021;96(1):24–30. doi:10.1097/ACM.0000000000003524

