## Supplement 2 for "Physician Facing AI Tools Show Distinct Failure Modes Under Structured Stress Testing"

**Data Sharing Statement**

**Data Availability**

All vignette prompts, model responses, and analysis datasets can be made available from the corresponding author upon reasonable request.

**Code Availability**

The R analysis code used in the study analysis and generating figures can be made available from the corresponding author upon reasonable request.
